# Impact of COVID-19 on the quality of life (QoL) of patients living with Sickle Cell Disorder (SCD) in Lagos, Nigeria

**DOI:** 10.1101/2021.09.17.21263748

**Authors:** Annette Akinsete, Michael Ottun, Hammed Adelabu, Larry Ajuwon, Jorden Veeneman

## Abstract

The study aimed to assess the impact of the COVID-19 pandemic on Quality of Life (QoL) in persons living with Sickle Cell Disorder (SCD) in Lagos, Nigeria and to determine how they coped during the pandemic, particularly during the period of total lockdown with the additional “SHIELDING” measures to which they had to adhere. Data was collected using a standardized protocol PedsQL, Sickle Cell Disease Module version.3.0 designed for youth within the ages of (13-18) years, (19-35) years and their parents/Guardian if underage. The survey captured data on patients’ pain impact, hurts, management, treatments, communication with their caregivers and their Guardian’s perception. The survey was performed online, or Face-to-Face/telephone interview if online was not possible. Contacts of patients and parents were obtained from the database of Sickle Cell Foundation Nigeria. A total of 105 (80 patients and 25 parents) participants responded to the survey. The age distribution of respondents was highest at 56 % in the age bracket of 13 - 18 years old. Pain crisis were very common amongst patients. The survey revealed that the type of treatment or care received at these times determined whether or not the patients visited the hospital when they had pain crises. In addition, as patients’ reports an increase in ill-treatment they experienced in the hands of health care givers, so did the fear of accessing treatment during the COVID pandemic. It was observed that the frequency of pain crises experienced by SCD patients was proportional to the patient’s quality of life (the higher the frequency of pains, the worse the QoL). As a follow-up, a more detailed study would be required, as this study was limited in the capturing of the demographics, sex and number of participants; Considering the number of persons living with SCD that visit the Sickle Cell Foundation Nigeria, (about 3,000 patients), the number of responses in this study was low (105). It is believed that a higher number of responses would have given more information about the Sickle Cell burden and the QoL of persons living with SCD in Lagos during the COVID-19 pandemic. Lagos was the epicentre of the COVID-19 pandemic in Nigeria.

## 1.0 Background of Study

COVID-19 is caused by the severe acute respiratory syndrome coronavirus 2 (SAR-CoV-2) of unknown origin, which was first reported in Wuhan, China in late 2019 (Ludwig and Zarbock, 2020) and declared a pandemic by the World Health Organisation (WHO) on March 11, 2020. The highly contagious nature of the virus has challenged public health systems across the globe with more than 177 million people affected and about 3.9 million deaths recorded in 216 countries as of June 20, 2021. While WHO and national health systems have created guidelines for management of the disease, effective protocols still are highly dependent on the strength of the national health systems (Akande and Akande,2020; WHO, 2021).

COVID-19 has altered the “***status quo***” of families, communities and the society because of enforcement of the “lockdown” strategy in many parts of the world, compelling people to stay at home - as a means of curtailing the spread of the virus. This affected health-seeking behavior of the citizenry, limiting access to health services during the heat of the pandemic. In Nigeria, a Sub-Saharan country, health care facilities did not accept patients because health workers feared contracting the virus. Particularly affected were those with non-communicable diseases (NCDs) who should routinely visit the clinic/hospital to receive care (Osseni, 2020).

The novel coronavirus can spread from infected patients through droplets and aerosols and its impact on infected individuals varies from mild to severe symptoms. Its notable impact on the respiratory system can lead to hypoxia, which can be fatal mostly in elderly patients (Ejaz et al., 2020; Hi et al., 2020). In addition, patients with pre-existing medical conditions have a higher risk, both for contracting the virus and for developing severe complications from the infection. This is particularly true for patients living with NCDs such as Hypertension, Diabetes, heart disease, cancer and sickle cell disorder (SCD), who often have a compromised respiratory system (John and John 2020).

Nigeria has the largest burden of SCD globally with an estimated 150,000 babies born annually with the condition (Akodu *et al*., 2013). It is therefore a major concern as patients living with SCD are immunocompromised and at risk of suffering from chronic or acute complications due to COVID which may require urgent hospitalisation and intensive care (John and John 2020). They describe the common presentation of COVID-19 infection in patients living with SCD. Symptoms include fever, shortness of breath, cough, and loss of smell and/or taste and in severe cases - pneumonia, multiple organ failure, sepsis and acute respiratory distress syndrome (ARDS).

Lagos State, Nigeria is considered the epicenter of the COVID-19 pandemic, recording up to one-third (18,177) of cases of infected subjects when compared with the other 35 states of the country and the Federal Capital Territory (which totals 54,463) (www.covid19.ncdc.gov.ng) accessed 2nd September, 2020.

Sadly, SCD and other NCDs have not gained much attention or garnered the political will that HIV/AIDS, tuberculosis and malaria have in the country. This presents tremendous challenges for addressing issues that relate to SCD in relation to COVID-19 (Heller *et al*., 2019; Thakur *et al*., 2020).

This is underlined by the fact that at the time of this publication, no cases of patients living with SCD who also contracted the coronavirus were reported in Nigeria.

### 1.1. Statement of Purpose

Curtailing the spread of COVID-19 is paramount in protecting vulnerable patients living with SCD and other NCDs. The debates rage on as to how to manage patients with COVID-19 best, especially patients with pre-existing medical conditions like SCD. The COVID-19 situation prevented patients living with SCD from accessing health care despite having health challenges ranging from pain crises to stroke, acute chest syndrome, leg ulceration and most importantly the psychological or mental health issues, which these patients have to contend with on a daily basis.

It is therefore important to know how patients with SCD coped with the effects of TOTAL LOCKDOWN and the additional “SHIELDING” measures that they had to adhere to during the COVID-19 pandemic.

### 1.2. Significance of Study

The stringent measures put in place to prevent patients living with SCD from contracting the coronavirus may lead to serious medical complications due to inability to access treatment and care through the usual means of routine hospital visits and follow-ups. The outcome of this survey will show real life challenges that patients living with SCD face during the COVID-19 pandemic and will offer suggestions on how to improve the Quality of Life (QoL) of patients living with SCD in these times.

### 1.3. Survey Objective

The objective of this survey was to assess the effect of the COVID-19 pandemic on the QoL of patients living with SCD in Lagos, Nigeria

## 2.0 METHODOLOGY

### 2.1. Study design and location

Participants’ contacts were identified from the Sickle Cell Foundation Nigeria Database from January 2018 to May 2020.

This survey was conducted online and Face-to-Face at the Sickle Cell Foundation Nigeria, located in Lagos State south west of Nigeria. The duration of the survey was three (3) months.

### 2.2 Data Collection

A standardized protocol PedsQL, Sickle Cell Disease Module version.3.0 was designed to collect data to assess the perceptions of parents and patients during the first lockdown due to COVID-19. The questionnaires were completed online, face-to-face or via telephone interview. Telephone and face-to-face interviews were performed for those who had challenges with mobile data to participate or who failed to complete the questionnaire online. The link to the questionnaire was sent to patients and their parents via SMS, WhatsApp and emails upon which feedback was received via the designed platform – RedCap (a secure web application for building and managing online surveys and databases developed by Vanderbilt University in 2004).

### 2.3 Inclusion Criteria

The survey was exclusively sent to patients living with SCD in Lagos, Nigeria, and who visited the Sickle Cell Foundation Nigeria. The age range of 13-18 years and 19-35 years was used and the survey was also sent to the Guardian (if applicable). Children below the age of 13, and patients without SCD were excluded from the survey.

### 2.4 Study Tool

We used a pre-validated questionnaire during the survey and links to subjects were provided by the Sickle Cell Foundation Nigeria. Information like patients’ pain impact, hurts, management, treatments and communication with their caregivers was collected and was used to assess the effect of COVID-19 on the QoL of participants.

### 2.5 Ethical Clearance

LUTH-HREC provided ethical clearance before survey commencement. The confidentiality of participants’ information was ensured during and after the survey by replacing patient identifiers with patient codes. Patients’ participation in the survey was voluntary and with the right to discontinue, without attracting any loss of rights and privileges to appropriate services at the center.

### 2.6 Data Analysis

SPSS 25.0 was used for data analysis. The descriptive statistics were used to present the demographics of the participants. The reliability of the questionnaire was tested using Cronbach’s Alpha. Linear regression analysis was also carried out to determine factors affecting the respondent’s fear during COVID as perceived by the participants.

### 2.6 Limitations

The limited sample size of this study was partly due to:

1. Limited literacy of patients,
2. Limited smartphone ownership, or
3. Limited access to Wi-Fi/internet.

Also, a general demographics questionnaire capturing gender for example was not used in this survey and should be added in future research.

In addition, we hope to conduct a Face-to-Face interview instead of by telephone (which has its limitations with respect to language barrier and may be expensive because of the cost of call credit). These we believe will further strengthen our statistical power and elucidate underlying needs not capture.

### 2.6. Funding

In-kind support was provided by RHIEOS Ventures.

## 3.0 RESULTS

### Descriptive statistics

208 respondents (which include patients living with SCD and their parents) participated in the survey, with 105 (80 patients and 25 parents) meeting the inclusion criteria. 103 participants were excluded due to; not meeting required age criteria and wrongly completing the questionnaire (affected respondents were not reachable). The eligible respondents participated via online questionnaire, telephone interview or Face-to Face questionnaire.

The majority of respondents was within the age bracket of 13-18 years old (56%). This corresponds well with the age reported by the parents being 72% within the age bracket of 13-18 years old respectively. (Table 1.0)

**Table 1.** Age distribution of respondents who had sickle cell disorder (SCD) or wards who had SCD

| Respondent living with SCD |  |  |
| --- | --- | --- |
| Age | Frequency | Percent |
| 13-18 | 42 | 56.0 |
| 19-35 | 33 | 37.7 |
| Missing age | 5 | 6.3 |
| Parents' ward age |  |  |
| 13-18 | 18 | 72.0 |
| 19-35 | 7 | 28.0 |

### 3.1 Reliability test

The reliability of the questions was tested using Cronbach’s Alpha. This showed a significant reliability with the least and highest scores of 0.531 and 0.831 respectively as shown in table 2.0. Hence, the questions in each construct when put together are a good measure of that construct.

**Table 2.0.** Reliability test of the questionnaire administered

| Variables | Respondents living with SCD |  | Parents of respondents living with SCD |  |
| --- | --- | --- | --- | --- |
|  | Cronbach's Alpha | No. of Items | Cronbach's Alpha | No. of Items |
| About my pain | .823 | 8 | .831 | 9 |
| Impact of my pain | .814 | 6 | .879 | 7 |
| About my worry | .849 | 7 | .782 | 7 |
| About my emotions | .756 | 2 | .754 | 2 |
| About my treatment | .675 | 6 | .530 | 6 |
| My communication about Sickle Cell | .790 | 7 | .709 | 5 |

Tables 3.0 and 4.0 (and figure 1 and 2) describe the participants’ responses mean based on the questions categorized into 8 parts. It is good to emphasise that the parents’ perceptions and that of their ward are very similar as shown in tables 3.0 and 4.0 respectively. To this end, only wards will be discussed within the scope of the survey.

**Table 3.0.**
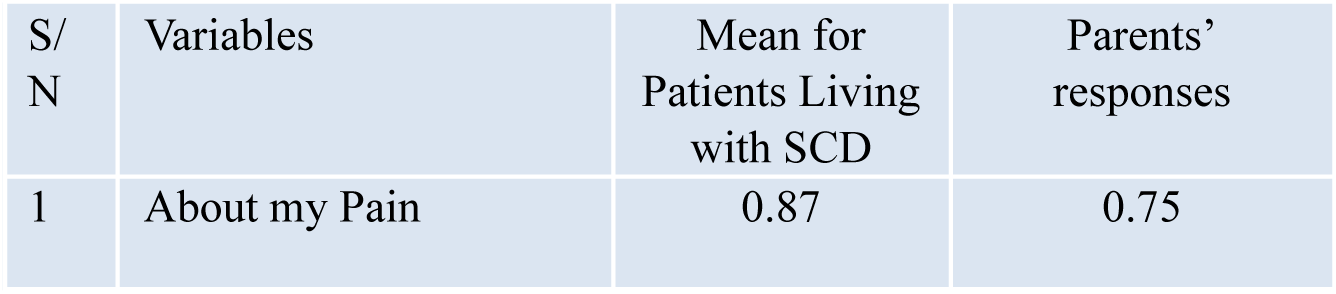
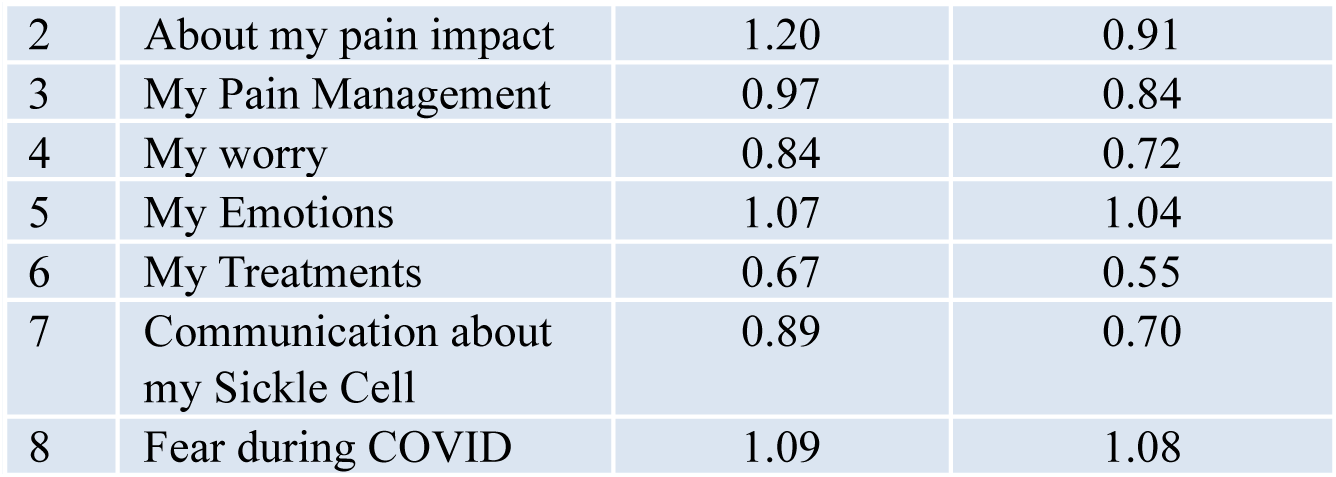
describing the mean score for the respondents.

**Table 4.0.**
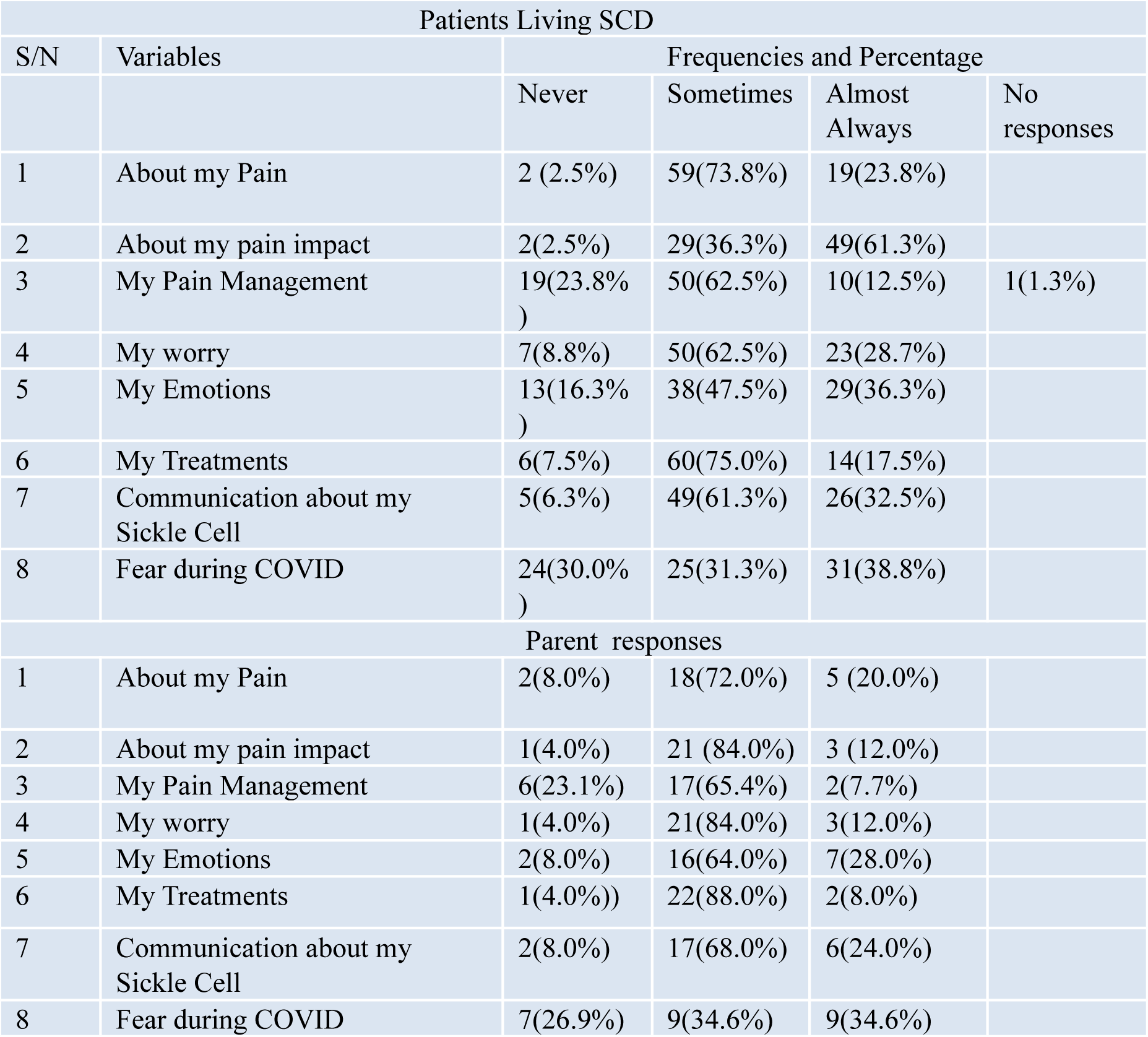
Frequencies and percentages of the Respondents’ responses.

| Patients Living SCD |  |  |  |  |  |
| --- | --- | --- | --- | --- | --- |
| S/N | Variables | Frequencies and Percentage |  |  |  |
|  |  | Never | Sometimes | Almost Always | No responses |
| 1 | About my Pain | 2 (2.5%) | 59(73.8%) | 19(23.8%) |  |
| 2 | About my pain impact | 2(2.5%) | 29(36.3%) | 49(61.3%) |  |
| 3 | My Pain Management | 19(23.8%) | 50(62.5%) | 10(12.5%) | 1(1.3%) |
| 4 | My worry | 7(8.8%) | 50(62.5%) | 23(28.7%) |  |
| 5 | My Emotions | 13(16.3%) | 38(47.5%) | 29(36.3%) |  |
| 6 | My Treatments | 6(7.5%) | 60(75.0%) | 14(17.5%) |  |
| 7 | Communication about my Sickle Cell | 5(6.3%) | 49(61.3%) | 26(32.5%) |  |
| 8 | Fear during COVID | 24(30.0%) | 25(31.3%) | 31(38.8%) |  |
| Parent responses |  |  |  |  |  |
| 1 | About my Pain | 2(8.0%) | 18(72.0%) | 5 (20.0%) |  |
| 2 | About my pain impact | 1(4.0%) | 21 (84.0%) | 3 (12.0%) |  |
| 3 | My Pain Management | 6(23.1%) | 17(65.4%) | 2(7.7%) |  |
| 4 | My worry | 1(4.0%) | 21(84.0%) | 3(12.0%) |  |
| 5 | My Emotions | 2(8.0%) | 16(64.0%) | 7(28.0%) |  |
| 6 | My Treatments | 1(4.0%)) | 22(88.0%) | 2(8.0%) |  |
| 7 | Communication about my Sickle Cell | 2(8.0%) | 17(68.0%) | 6(24.0%) |  |
| 8 | Fear during COVID | 7(26.9%) | 9(34.6%) | 9(34.6%) |  |

**Fig. 1.**
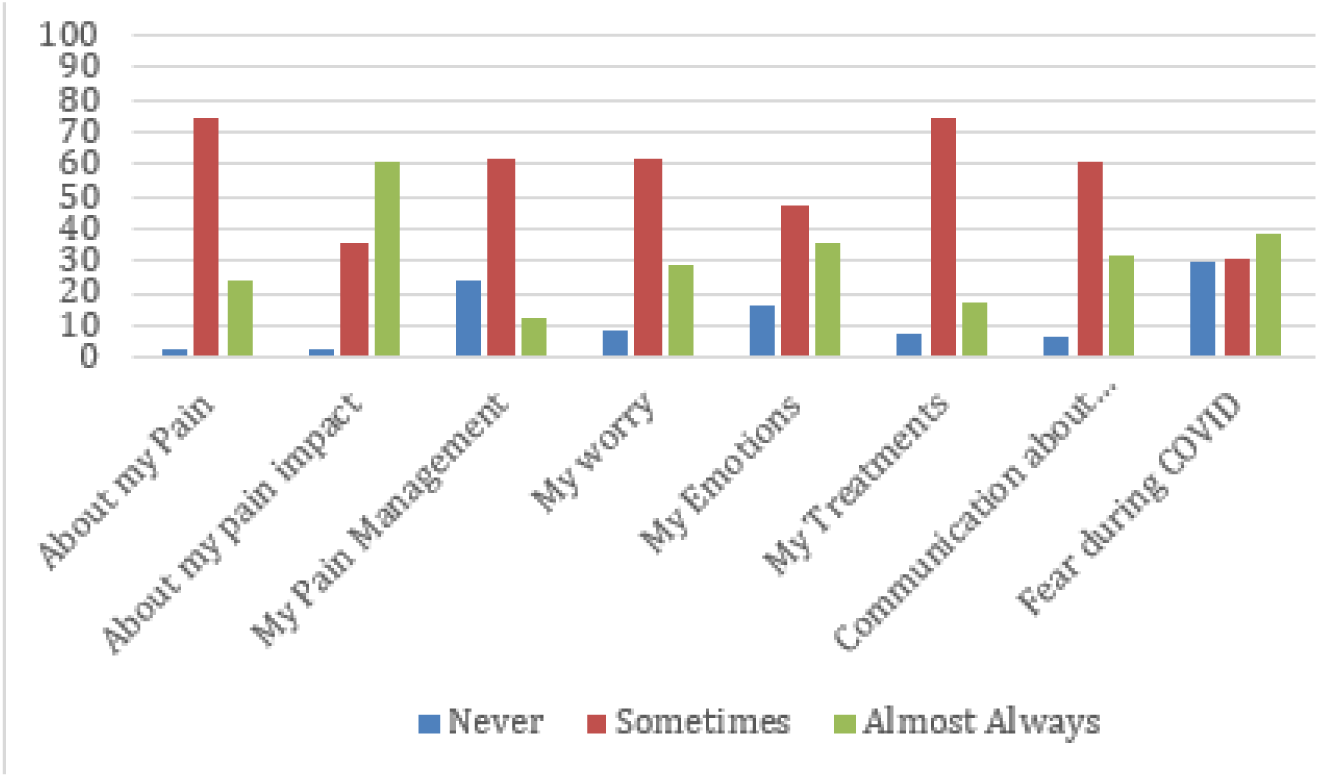
shows the summary of the wards’ responses

**Fig. 2.**
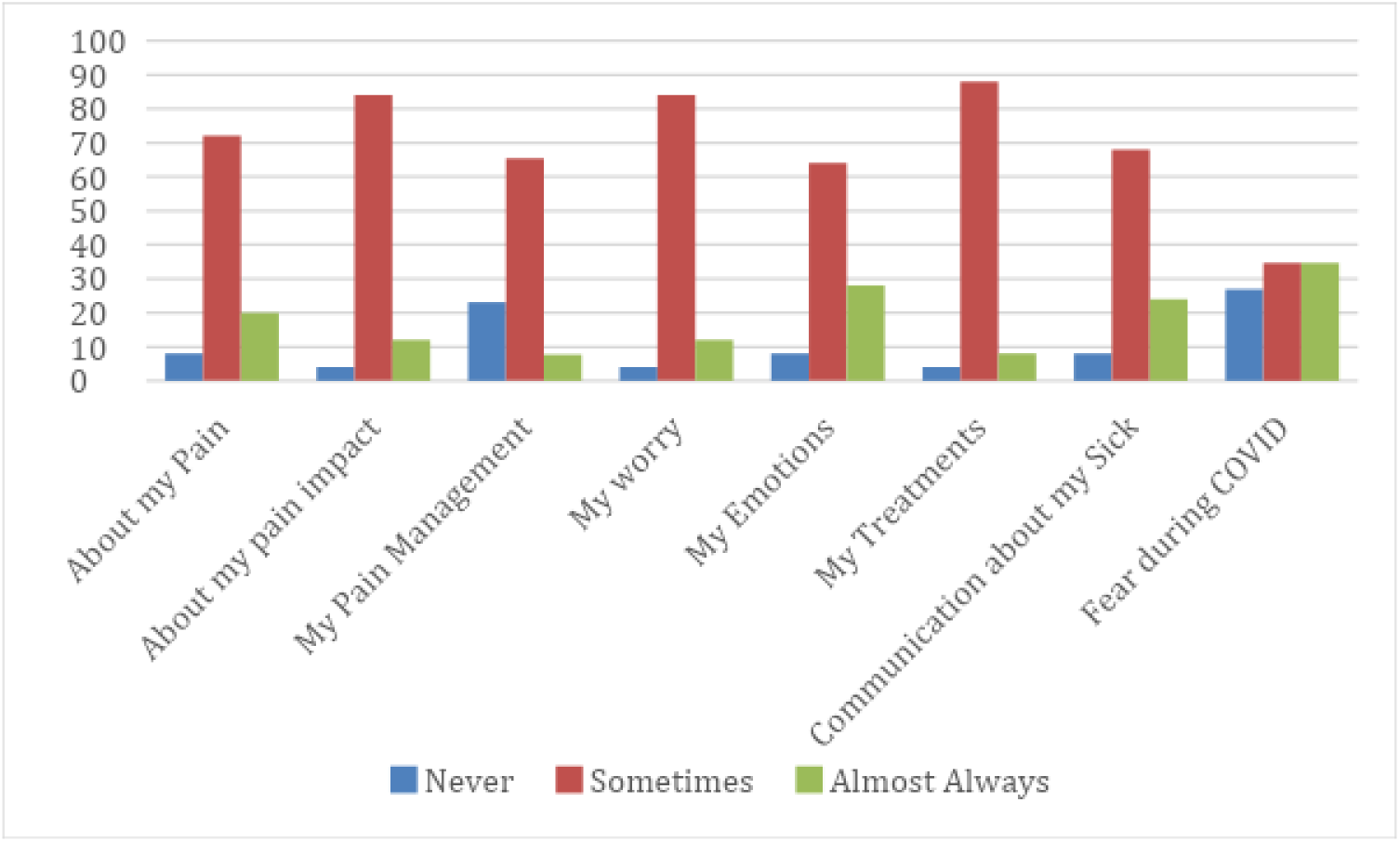
shows the summary of parents’ perception of their ward.

### 3.2 About “my pain” and “Impact of pain”

The pain frequency in all parts of their body experienced amongst wards living with SCD was shown to be highest with 97.6% and with a mean score of (0.87). The impact of pain was highest in wards with frequency of 29 (96.0%) and a mean score of (1.20).

### 3.3 Pain management

Pain management showed a mean of 0.97 and a frequency of 76.0% of wards who sometimes find it hard to manage their pain.

### 3.4 About “my worry”

Responses of the wards’ worries revealed that a higher percentage of 91.2% (with a mean of 0.84) worried about their pain and the fear being hospitalised as presented in table 3.0 and 4.0

### 3.5 About “my treatments”

Tables 3.0 and 4.0 show responses about treatments, most of the wards (92.5% and a mean of 0.67) sometimes did not remember to take their medication on account of it making them sleepy, the taste of the drug(s), or even forgetting to take them.

### 3.6 About “my emotions”

The wards’ emotions showed that 83.8% (as shown in table 4.0) of wards are very angry about their genotype and pain.

### 3.7 Communication about my SCD

The responses about communication showed that 93.8% of the wards find it difficult to speak with their caregiver or speak to anyone about their pain or their genotype as seen in table 3.0 and 4.0 respectively.

### 3.8 Fear of restriction due to COVID-19

The data also revealed that the majority of the wards (70.1 %) were afraid to attend clinic during the pandemic due to fear of restriction.

### 3.9 Linear Regression Analysis

The factor affecting the respondents’ fear during COVID-19 pandemic, as perceived by the wards and their parents, was assessed using linear regression analysis,. Of the variables measured, the wards’ perception of the nature of treatment they received (i.e. attitude of health caregivers towards them) was the only significant factor affecting their fear of receiving treatment during the peak of the COVID-19 pandemic. Table 5.0 showed that as the measure of the wards’ report on the treatment received (i.e. negative attitude of health care givers) increased, the fear of getting treatment during COVID was likely to increase by a factor of 1.043, as shown in Table 5.

**Table 5.** Linear regression analysis showing the factors affecting the respondent’s fear during COVID as perceived by the child

**a. Model Summary**
| Model | R | R Square | Adjusted R Square | Std. Error of the Estimate |
| --- | --- | --- | --- | --- |
| 1 | .590<sup>a</sup> | .348 | .283 | .707 |

a. Predictors: (Constant), My communication about my sickle cell, About my pain, my emotions, My pain management, My worrying, My treatment, Impact of my pain

**b. ANOVA<sup>a</sup>**
| Model |  | Sum of Squares | df | Mean Square | F | Sig. |
| --- | --- | --- | --- | --- | --- | --- |
| 1 | Regression | 18.900 | 7 | 2.700 | 5.403 | .000<sup>b</sup> |
|  | Residual | 35.480 | 71 | .500 |  |  |
|  | Total | 54.380 | 78 |  |  |  |

**c. Coefficients<sup>a</sup>**
| Coefficients<sup>a</sup> |  |  |  |  |  |  |
| --- | --- | --- | --- | --- | --- | --- |
| Model |  | Unstandardized Coefficients |  | Standardized Coefficients | T | Sig. |
|  |  | B | Std. Error | Beta |  |  |
| 1 | (Constant) | .063 | .236 |  | .268 | .790 |
|  | About my pain | -.212 | .311 | -.096 | -.683 | .497 |
|  | Impact of my pain | .398 | .290 | .219 | 1.374 | .174 |
|  | My pain management | .143 | .171 | .102 | .836 | .406 |
|  | My worrying | .099 | .253 | .055 | .392 | .696 |
|  | my emotions | -.155 | .158 | -.127 | -.978 | .331 |
|  | My treatment | 1.043 | .302 | .489 | 3.451 | .001 |
|  | My communication about my sickle cell | -.010 | .227 | -.006 | -.043 | .966 |

a. Dependent Variable: Fear during COVID

However, the analysis of parents’ perception of the wards’ emotion, the wards’ treatment, the wards’ communication of their treatment and the report of how the pain was managed had no significant effect on the fear of getting treatment during COVID (data not shown). This is probably due to the smaller sample size.

## 4.0 DISCUSSION AND CONCLUSION

The survey reveals that pain due to SCD in children or adults is common and a major challenge when it comes to its management. Pain in SCD could also vary with respect to its severity and as such there is a need for medical attention, in most cases with hospitalization, especially in children (Borhade and Kondamudi, 2021). Study by Lakkakula *et al*., 2018 showed that pain management in children living with SCD can be complex and difficult and should require some kind of pain assessment.

Pain, which is common amongst persons living with SCD, can be worrisome and can be a nightmare not only to the person but also to their parents. This is especially true in cases where there is poor pain management due to lack of funds, access to the hospital, unpalatable experiences with health workers, fear for restriction from the hospital during the COVID lockdown, or lack of experts or caregivers to get adequate treatment.

The frequency of pain crises experienced by SCD patients is proportional to the patients’ quality of life (the higher the frequency of pains, the worse the QoL) as discussed by Shah *et al*., (2019).

This survey revealed that the kind of treatment or care (referencing the attitude of health workers) received determines whether subjects visited the hospital whenever they had pain crises. The delay in being attended to at the hospital during an emergency or routine appointment, the attitude and communication from caregivers were factors that deterred parents and patients from attending the clinic. Taber *et al*., 2015. Study by Rees *et al*., 2003 also emphasised the need for continual education and selfless interest for nurses caring for SCD patients during their pain crises.

Sadly, the nature of the treatment and care or the experience of pain management during crises may lead to some notable detrimental effects in patients with SCD - as emotional, behavioral, attendance at clinic, affective and psychological responses as deduced from the survey and as reported in a study by Dunlop and Bennett in 2014.

The perception of parents about their childs’ emotion and pain management could be a determining factor for their child’s access to treatment. The health providers, especially the nurses and counselors, do have a key role to play in communicating at what point a parent or patient ought to seek medical attention. Currently, most of the pain crises are managed at home until the pain is unbearable (Leger *et al*., 2018; Tanabe *et al*., 2020).

The survey also revealed that it is common for people living with SCD not to disclose their pain or genotype to their caregiver or to anyone and this may be attributed to them not wanting to be stigmatized. As studies by *Leger et al*., 2018 reported the impact of stigma on behaviour in individuals which presented as psychological stress, depression, late diagnosis and inadequate management.

Therefore, there is a need for adequate awareness by the government and NGOs to sensitise and address the issue of stigmatisation amongst SCD patients. There is also a need for greater political will and government’s effort in drawing attention to SCD, enacting policies and legislation around affordable services, newborn screening, provision of drugs, Genetic counselling just as has been done in the past to reduce the burden of HIV/AIDS (Awofala and Ogundele, 2018).

Sickle Cell Disorder and other non-communicable diseases should be given more attention, as statistics during the pandemic has shown that patients with underlying conditions gives a twofold increase in risk of having severe COVID-19 complications.

The preventive measures like face masking, washing and sanitizing the hands, and zero tolerance to crowded areas are there for the whole world. But it is imperative having dedicated clinics with trained experts (especially doctors, genetic counsellors and other relevant health workers) for better treatments, education and care in order to reduce not just their risk to COVID complications, but also improve the quality of life (QoL) in the sickle cell community.

Going forward, there is still a need for a more detailed study, as this study is limited in the number of respondents and the number of patients living with sickle cell that visit the Sickle Cell Foundation (about 3,000 patients,). More and more complete responses may give information about the Sickle burden and the QoL of the patients during the pandemic in Lagos State which was the epicentre of the COVID-19 pandemic in Nigeria.

## Data Availability

Survey response are available in the Sickle Cell Foundation Registry database.

https://www.sicklecellfoundation.com/research-survey/

